# Tobacco Smoking and Associated Factors among In-school Adolescents in Vietnam in 2013, 2019

**DOI:** 10.1101/2021.03.28.21254501

**Authors:** Hoang Van Minh, Khuong Quynh Long, Do Van Vuong, Nguyen Manh Hung, Kidong Park, Momoe Takeuchi, Mina Kashiwabara, Nguyen Tuan Lam, Pham Thi Quynh Nga, Le Phuong Anh, Le Van Tuan, Tran Quoc Bao, Le Duong Minh Anh, Tran Thi Tuyet Hanh

## Abstract

Tobacco smoking is one of the most dangerous risk behaviors, leading to many adverse human health consequences. The aims of this study is to estimate the prevalence of tobacco smoking and related factor among adolescents aged 13–17 years in Vietnam. The data were from two rounds of the Vietnam Global School-based Student Health Survey (GSHS) that is the nationally representative survey conducted in 2013 and 2019. The logistic regressions were carried out to identify factors associated with tobacco smoking among study participants. We found the prevalence of current smoking (water pipe and cigarettes) reduced significantly from 5.4% (95% CI: 4.0–7.2) in 2013 to 2.8% (95% CI: 2.2–3.6) in 2019. In 2019, 2.6% of students used electronic cigarette products in the last 30 days. Factors associated with higher odds of current smoking status included study year, gender, parental monitoring, loneliness, suicide attempt, sexual intercourse, truancy, alcohol drinking. Similar patterns were found in e-cigarette use. Smoking among adolescents in Vietnam reduced between 2013 and 2019. Further follow-up studies are needed to confirm the causal factors of the reduction and e-cigarettes use.

## Introduction

Tobacco smoking is one among the most dangerous risk behaviors, leading to many adverse human health consequences, such as cancers, cardiovascular diseases, chronic pulmonary diseases and many other types of health problems [1, 2]. Tobacco smoking is related to more than 8 million cases of death annually and this figure will continue to increase in the coming time [1]. Around 80% of 1.3 billion smokers are living in lower-middle income countries (LMIC), where tobacco-related burden of disease is the most severe [2].

Compelling research evidence showed that the age of first tobacco smoking is in the trend of decreasing [3-5]. More than 50% of current smokers start to smoke since adolescent [2, 6]. In fact, according to the Global Youth Tobacco Survey conducted in 61 countries between 2012 and 2015, the median proportion of current smokers among students aged 13-15 years old was 10.7% [7, 8]. Adolescent smoking before 18 year old tends to form a habit and should be harder to quit than the others [9].

Vietnam is one of the 15 countries with highest proportion of male smokers in the world. The prevalence of current smoking among adults aged 15 years old in Vietnam in 2015 was 22.5% (45.3% among men, 1.1% among women) [10]. Approximately 75,000 people dying each year due to complications caused by smoking [11]. In 2011, the total economic cost related to smoking was around US$ 1.2 billion, equivalent to 5.76% of the national healthcare budget [12]. Although the prevalence of smoking among adolescents in Vietnam seems to be still not high, i.e. 3.5% (6.3% among boys and 0.9%) in 2014 among the children aged 13-15 years old [13], the issues of youth smoking in Vietnam should not be underestimated because tobacco companies are trying to expand cigarette markets by promoting cigarettes to adolescents [9]. Also, youth smokers are three times more likely than non-smokers to use alcohol, eight times more likely to use marijuana, and 22 times more likely to use cocaine. Smoking is associated with a host of other risky behaviors, such as fighting and engaging in unprotected sex [9].

In 2001, to support the tobacco prevention and control efforts, WHO in collaboration with UNAIDS, UNESCO, UNICEF, and with technical assistance from the U.S. CDC initiated the development of the Global School-based Student Health Survey (GSHS). GSHS is a collaborative surveillance project designed to help countries measure and assess the behavioral risk factors and protective factors in 10 key areas among young people aged 13 to 17 years. The GSHS is a relatively low-cost school-based survey, which uses a self-administered questionnaire to obtain data on young people’s health behavior and protective factors related to the leading causes of morbidity and mortality among children and adults worldwide [14]. In Vietnam, the GSHS was conducted in 2013 and 2019 to provide data on health behaviors and protective factors among students. aiming to help developing priorities, establish programs, and advocate for resources for school health and youth health programs and policies [15, 16]. In this paper, using the data from the Vietnam GSHS 2013 and 2019, we aim to describe the prevalence of tobacco smoking and associated factors among students aged 13-17 years.

## Methods

### Data source

The Vietnam GSHS 2013 and 2019 used the standardized protocol of WHO with technical support from the US CDC, which was designed to collect data from students in developing countries [17]. The Vietnam GSHS employed a two-stage cluster sampling design to produce a nationally representative sample of school students aged 13-17. In the first stage, schools were selected based on a probability proportional to enrolment size. In the second stage, in each selected school, classes were chosen using the simple random sampling technique. The final sample for GSHS 2013 was 3331 students from 13 cities/provinces (response rate was 96.0%), while the sample size for GSHS 2019 was 7796 students from 20 cities/provinces (response rate was 93.5%).

### Questionnaire development

In the Vietnam GSHS 2013 survey, the GSHS standardized questionnaire in English [17] was translated into Vietnamese using the forward and backward translation. The 2013 translated questionnaire was pilot tested with 50 students before official use. In the GSHS 2019, we used the 2013 version with some updates as per the 2019 original English version. We also used relevant indicators from the 25 Global Non-Communicable Diseases and Healthy Vietnam [18] to adapt into the Vietnam GSHS 2019 questionnaire. The 2019 questionnaire was pilot tested with 120 students aged 13–17 years for the comprehension of questions, the appropriateness of language, and checking for any logical issues. The pilot data were reviewed by senior researchers to suggest modifications to the translation to improve clarity.

### Data collection

In each selected class, a homeroom teacher supported us to explain to students about the study. We provided information about the Vietnam GSHS and consent forms for the students and requested them to share these documents with parents for signing as an agreement to participate in the study. The following day, we administered GSHS questionnaire among students who had consent from their parents and were willing to participate in the study. Data collectors, who were medical or public health students, were trained on both GSHS questionnaire and data collection procedure, took main responsibilities for the data collection process. Students answered the self-administered questionnaire on separate, computer scanable answer sheets, during one classroom period. All the procedure of data collection was following WHO’s guidance and templates.

### Measurements

In this paper, the dependent variable was current smoking status (smoking in the past 30 days). The independent variables included demographic characteristics, including gender (male/female) and age (from 13 to 17); parental factors, including parental monitoring (parents most of the time/always checked homework or know what their children do in their free time – yes/no), parental understanding (parents most of the time/always understood problems/worries or gave advice/guidance – yes/no), and parental respect (parents most of the time/always respect their children or their personal space – yes/no); have close friend (students who had any close friends – yes/no); mental health, including loneliness (students who most of the time/always felt lonely during the past 12 months – yes/no) and suicide attempt (students who seriously considered attempting suicide in the past 12 months – yes/no); violence (students who were physically attacked in the past 12 months – yes/no); bullying (students who were bullied at school in the past 30 days – yes/no); sexual intercourse (students who have ever had sexual intercourse – yes/no); truancy (students who missed any days without permission during the past 30 days – yes/no); risk behaviors, including drinking alcohol (students who drank at least 1 drink in the past 30 days – yes/no), sedentary behaviors (student who spent three or more hours a day to do these activities – yes/no), and low fruit/vegetable intake (students who ate less than 5 time fruits/vegetables per day in the past 30 days – yes/no). We also assessed the smoking environment of students defined as students who had seen/contacted with smokers in the past 7 days (yes/no). The questions used to identify smoking status and other classification of statuses are described in **Table1**.

**Table 1:**
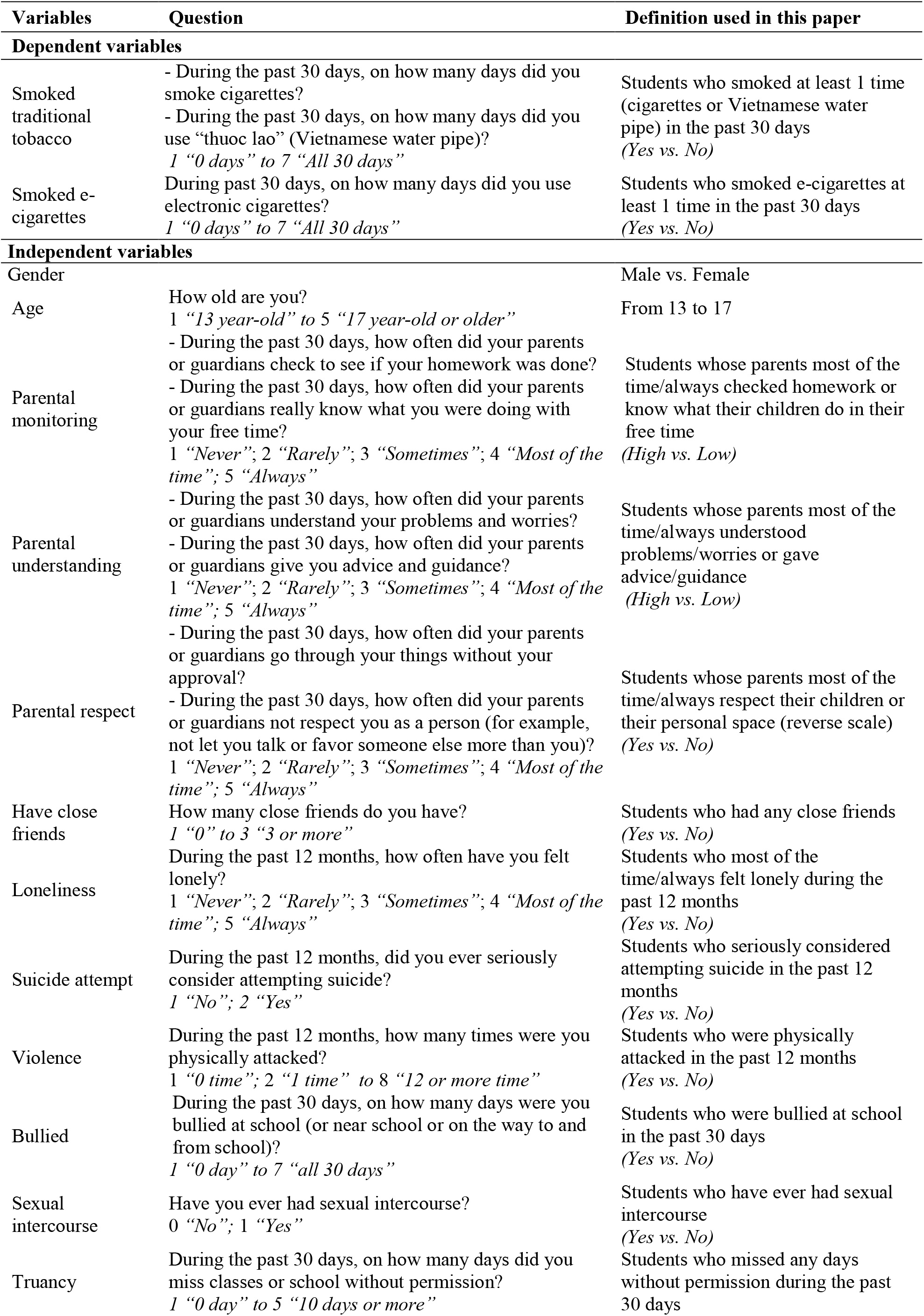
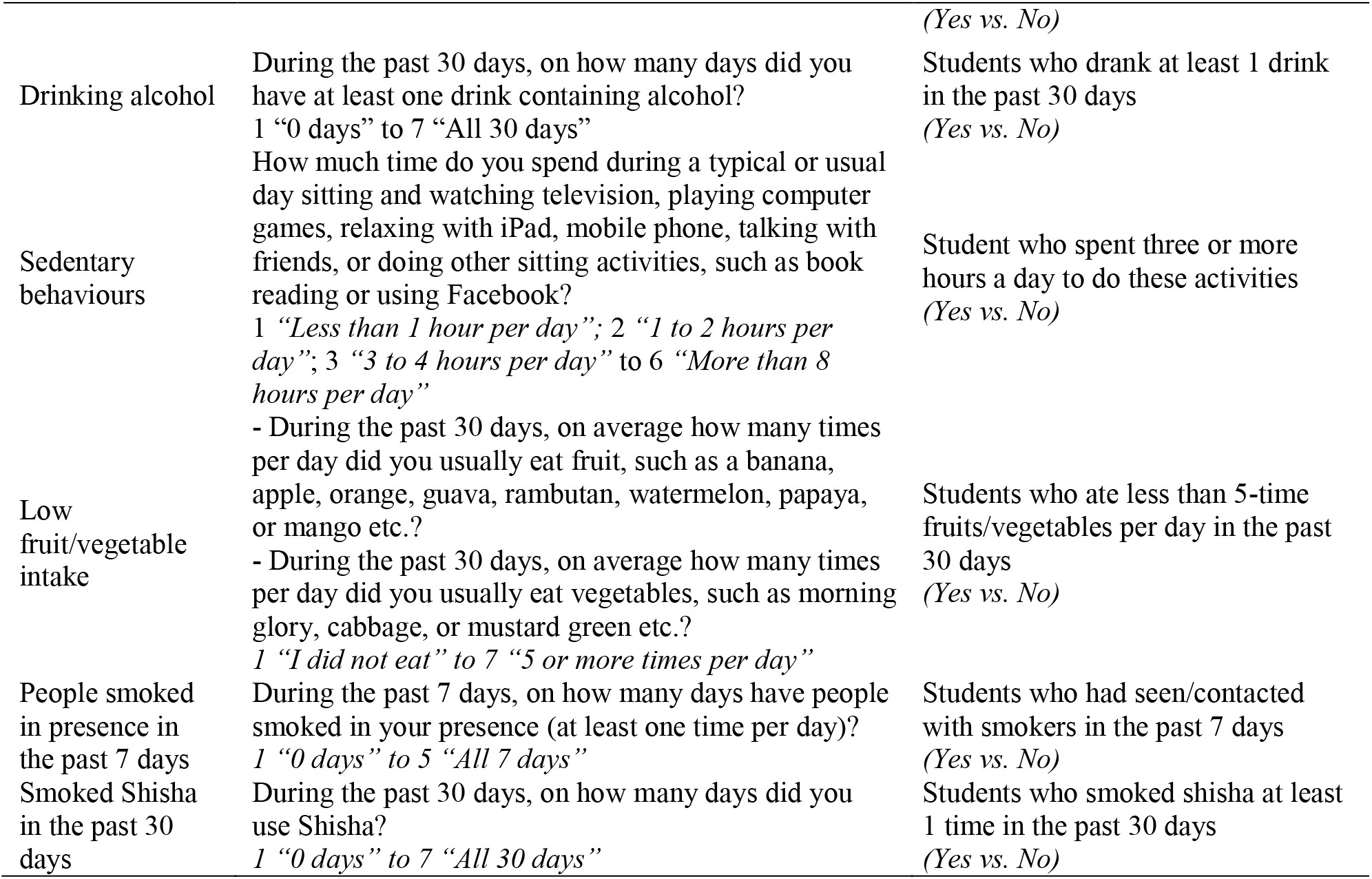
Definitions of dependent and independent variables analysed.

### Data processing and analysis

Data entry was conducted using a scanning device and then transformed into the data frame using Stata software. The data set collected were cleaned and edited for inconsistencies, following the guideline for data users published at the CDC website. A weighting factor was applied to each student record to reflect the likelihood of sampling each student and to reduce bias by compensating for different patterns of nonresponse. The weight used for estimation was given by.

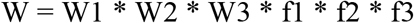

In which:

W1 is the inverse of the probability of selecting the province

W2 is the inverse of the probability of selecting the school

W3 is the inverse of the probability of selecting the classroom within the school

f1 is a school level nonresponse adjustment factor calculated by school.

f2 is a student level nonresponse adjustment factor calculated by class size category. The factor was calculated in terms of class enrolment instead of number of classes.

f3 is a post-stratification adjustment factor calculated by rural/urban and grade.

In GHSH 2019, among 7796 students, 106 self-identified as other-gender. Due to the small sample size of the other-gender group, and to be consistent with GSHS 2013, we included male and female students (n = 7690) in the analysis. Logistic regressions were used to identify factors related to current cigarettes and e-cigarettes smoking. All analyses were conducted using Stata v16 with survey package *“svy”* to take into account the complex sample design (strata, cluster and weights).

### Consent and ethical consideration

This survey was approved by the Ethics Committee of Hanoi University of Public Health (IRB No 421/2019/YTCC-HD3 dated 06^th^ August 2019). The study also received supports from Ministry of Health, Ministry of Education and Training, Provincial Department of Education and Training, and selected schools. All students participating in this survey were explained the objectives of the study. Survey procedures were designed to protect student privacy by allowing for anonymous and voluntary participation. Specifically, their participations were completely voluntary, which was clearly indicated in the Consent Form for Participation in the Research. The students had the right to withdraw from the study or refuse to answer any specific questions in the questionnaire without any consequences. All personal information of school students was kept confidential, and all information was encrypted.

## Results

### Characteristics of the study students

The characteristics of the study students are presented in **Table 2**. The patterns of parental monitoring, having close friends, reporting loneliness, suicide attempt, sexual intercourse and sedentary behaviors were quite similar in both study years. The percentages of students who reported having parental understanding parental respect were higher in the GSHS 2019. The proportions of student who experienced violence, bully, truancy and drinking alcohol in the past 30 days were lower in GSHS 2019.

**Table 2:** Participants’ characteristics.

| Characteristics | 2013 |  | 2019 |  |
| --- | --- | --- | --- | --- |
|  | n | Weighted % | n | Weighted % |
| **N** | **3331** |  | **7690** |  |
| **Gender** |  |  |  |  |
| Male | 1,557 | 46.9 | 3,572 | 46.0 |
| Female | 1,765 | 53.1 | 4,118 | 54.0 |
| **Age** |  |  |  |  |
| 13 | 897 | 21.7 | 1,024 | 16.7 |
| 14 | 858 | 22.5 | 1,523 | 30.4 |
| 15 | 542 | 19.6 | 1,672 | 19.9 |
| 16 | 769 | 23.9 | 1,646 | 15.9 |
| 17 | 264 | 12.3 | 1,825 | 17.1 |
| **Parental monitoring** |  |  |  |  |
| Low | 1,664 | 52.2 | 4,252 | 52.3 |
| High | 1,662 | 47.8 | 3,410 | 47.7 |
| **Parental understanding** |  |  |  |  |
| Low | 2,283 | 69.5 | 3,586 | 44.2 |
| High | 1,038 | 30.5 | 4,102 | 55.8 |
| **Parental respect** |  |  |  |  |
| Low | 1,209 | 36.8 | 1,176 | 14.3 |
| High | 2,070 | 63.2 | 6,451 | 85.7 |
| **Have close friends** |  |  |  |  |
| No | 173 | 5.5 | 778 | 8.8 |
| Yes | 3,138 | 94.5 | 6,816 | 91.2 |
| **Loneliness** |  |  |  |  |
| No | 2,910 | 88.5 | 6,589 | 87.7 |
| Yes | 367 | 11.5 | 1,098 | 12.3 |
| **Suicide attempt** |  |  |  |  |
| No | 2,750 | 83.1 | 6,333 | 84.6 |
| Yes | 539 | 16.9 | 1,347 | 15.4 |
| **Violence** |  |  |  |  |
| No | 2,582 | 79.0 | 6,885 | 89.6 |
| Yes | 734 | 21.0 | 802 | 10.4 |
| **Bullied** |  |  |  |  |
| No | 2,456 | 77.3 | 7,258 | 94.5 |
| Yes | 744 | 22.7 | 427 | 5.5 |
| **Sexual intercourse** |  |  |  |  |
| No | 3,002 | 93.5 | 7,188 | 94.9 |
| Yes | 187 | 6.5 | 459 | 5.1 |
| **Truancy** |  |  |  |  |
| No | 2,725 | 80.6 | 6,307 | 84.9 |
| Yes | 598 | 19.4 | 1,191 | 15.1 |
| **Drinking alcohol in the past 30 days** |  |  |  |  |
| No | 2,476 | 75.1 | 5,781 | 77.9 |
| Yes | 722 | 24.9 | 1,864 | 22.1 |
| **Sedentary behaviors** |  |  |  |  |
| No | 1,968 | 58.0 | 3,942 | 57.1 |
| Yes | 1,349 | 42.0 | 3,683 | 42.9 |
| **Low fruit/vegetables intake** |  |  |  |  |
| No | 693 | 20.5 | 370 | 4.6 |
| Yes | 2,603 | 79.5 | 7,295 | 95.4 |

### Pattern of current tobacco smoking among the study students

As shown in **Table 3**, the prevalence of current tobacco smoking (both water pipe and cigarettes) among the study students in 2013 and 2019 were 5.4% (95% CI: 4.0–7.2) and 2.8% (95% CI: 2.2–3.6). The difference was statistically significant. Significant decreases in the prevalence of smoking among the study students from 2013 to 2019 were found for both cigarette smoking and water pipe. In 2019, 2.6% of students reported that they used electronic cigarette products in the last 30 days.

**Table 3:** Smoking prevalence and trend from 2013 to 2019.

| Characteristics | 2013 |  | 2019 |  |
| --- | --- | --- | --- | --- |
|  | % | 95% CI | % | 95% CI |
| Have ever smoked | 12.1 | 9.8–15.0 | 8.2 | 7.0–9.6 |
| Smoked cigarettes in the past 30 days | 4.7 | 3.5–6.3 | 2.6 | 2.0–3.4 |
| Smoked water pipe in the past 30 days | 2.4 | 1.5–3.8 | 1.0 | 0.7–1.4 |
| Traditional tobacco smoking (cigarettes and water pipe) in the past 30 days | 5.4 | 4.0–7.2 | 2.8 | 2.2–3.6 |
| People smoked in presence in the past 7 days | 75.8 | 73.4–78.0 | 66.0 | 63.8–68.2 |
| Smoked Shisha in the past 30 days | n/a | n/a | 1.3 | 0.93–1.72 |
| Smoked e–cigarettes in the past 30 days | n/a | n/a | 2.6 | 1.9–3.3 |

The prevalence of current traditional tobacco smoking and e-cigarettes smoking (and 95% CI) among the study students by their characteristics (among all study students and the male students) are shown in **Supplemental Table 1** and **Supplemental Table 2**.

### Factors associated with traditional tobacco smoking among the study students

Table 4 shows the results of logistic regression analyses of factors associated with traditional tobacco smoking status among the study students. Among all students, after controlling for other independent variables in the model, the statistically significant factors associated with current smoking status were: 1) study year: lower in 2019 (OR = 0.66); 2) gender: higher among male student (OR = 4.20); 3) parental monitoring: lower in the group with high level of parental monitoring (OR = 0.57); 4) loneliness: higher among those who felt loneliness (OR = 1.57); 5) suicide attempt: higher among those who had suicide attempt (OR = 1.71); 6) sexual intercourse: higher among those who had sexual intercourse (OR = 4.56); 7) truancy: higher among those who reported truancy (OR = 2.23); 8) alcohol drinking: higher among those who drank alcohol in the past 30 days (OR = 4.17). Among male students, after controlling for other independent variables in the model, statistically significant association was found with age, with higher odds of tobacco smoking among older students (OR = 2.0-2.3). All associated factors found in the model for all students remained significant in the model for males, excepting for study year, loneliness, and suicide attempt.

**Table 4:** Factors related to traditional tobacco smoking among students aged 13–17 in Vietnam.

| Factor | All students |  | Male students |  |
| --- | --- | --- | --- | --- |
|  | OR | 95% CI | OR | 95% CI |
| **Year, (Ref: 2013)** |  |  |  |  |
| 2019 | **0.66\*** | **0.44-0.99** | 0.63 | 0.39-1.00 |
| **Gender, (Ref: Female)** |  |  |  |  |
| Male | **4.25\*\*\*** | **2.56-7.04** | -- | -- |
| **Age, (Ref: 13)** |  |  |  |  |
| 14 | 1.19 | 0.71-2.00 | 1.40 | 0.63-3.11 |
| 15 | 1.27 | 0.85-1.92 | **2.00\*** | **1.16-3.44** |
| 16 | 1.41 | 0.86-2.30 | **2.30\*\*** | **1.24-4.27** |
| 17 | 1.45 | 0.93-2.25 | **2.30\*** | **1.18-4.48** |
| **Parental monitoring, (Ref: Low)** |  |  |  |  |
| High | **0.57\*** | **0.37-0.88** | **0.58\*** | **0.34-0.97** |
| **Parental understanding, (Ref: Low)** |  |  |  |  |
| High | 0.90 | 0.63-1.30 | 0.97 | 0.62-1.51 |
| **Parental respect, (Ref: Low)** |  |  |  |  |
| High | 0.73 | 0.51-1.04 | 0.84 | 0.57-1.24 |
| **Have close friends, (Ref: No)** |  |  |  |  |
| Yes | 1.28 | 0.73-2.23 | 1.19 | 0.61-2.31 |
| **Loneliness, (Ref: No)** |  |  |  |  |
| Yes | **1.57\*** | **1.10-2.25** | 1.37 | 0.82-2.26 |
| **Suicide attempt, (Ref: No)** |  |  |  |  |
| Yes | **1.71\*** | **1.10-2.67** | 1.61 | 0.84-3.06 |
| **Violence, (Ref: No)** |  |  |  |  |
| Yes | 1.31 | 0.80-2.17 | 1.36 | 0.77-2.38 |
| **Bullied, (Ref: No)** |  |  |  |  |
| Yes | 0.82 | 0.50-1.33 | 0.89 | 0.50-1.59 |
| **Sexual intercourse, (Ref: No)** |  |  |  |  |
| Yes | **4.56\*\*\*** | **3.17-6.57** | **4.57\*\*\*** | **3.13-6.66** |
| **Truancy, (Ref: No)** |  |  |  |  |
| Yes | **2.23\*\*** | **1.38-3.61** | **2.31\*\*** | **1.29-4.14** |
| **Drinking alcohol in the past 30 days, (Ref: No)** |  |  |  |  |
| Yes | **4.17\*\*\*** | **2.78-6.24** | **3.79\*\*\*** | **2.31-6.22** |
| **Sedentary behaviors, (Ref: No)** |  |  |  |  |
| Yes | 0.98 | 0.71-1.36 | 0.86 | 0.61-1.22 |
| **Low fruit/vegetables intake, (Ref: No)** |  |  |  |  |
| Yes | 1.11 | 0.73-1.67 | 1.28 | 0.71-2.31 |

\* $p < 0.05$ ; \*\* $p < 0.01$ ; \*\*\* $p < 0.001$

### Factors associated with e- cigarettes smoking among the study students

Factors associated with e-cigarette use among students in 2019 are presented in **Table 5**. We found that being male, attempting suicide in the past 12 months, having sexual intercourse, being bullied, truancy, drinking alcohol, and spending three or more hours on sedentary behaviors were significant factors increasing the odds of using e-cigarettes among students. Especially, students who reported drinking alcohol in the past 30 days were five times more likely to be an e-cigarette user (OR = 5.38, 95% CI: 3.75-7.71). However, having high parental monitoring and respect decreased the odds of being e-cigarette users by 44% and 36% among students, respectively. All these associated factors remained significant in the regression model for male students, excepting parental respect, and truancy. Age, parental understanding, loneliness, violence suffering, and fruit/vegetable intake were found to be not significantly associated with e-cigarette use among students.

**Table 5:** Factors related to e-cigarettes smoking among students aged 13–17 in Vietnam.

| Factor | All students |  | Male students |  |
| --- | --- | --- | --- | --- |
|  | OR | 95% CI | OR | 95% CI |
| **Gender, (Ref: Female)** |  |  |  |  |
| Male | **2.09\*\*** | **1.28-3.42** |  |  |
| **Age, (Ref: 13)** |  |  |  |  |
| 14 | 0.56 | 0.22-1.44 | 0.57 | 0.19-1.72 |
| 15 | 0.46 | 0.17-1.25 | 0.67 | 0.21-2.13 |
| 16 | 0.42 | 0.14-1.23 | 0.56 | 0.18-1.73 |
| 17 | 0.64 | 0.21-2.00 | 1.08 | 0.33-3.50 |
| **Parental monitoring, (Ref: Low)** |  |  |  |  |
| High | **0.56\*\*** | **0.40-0.78** | **0.54\*\*** | **0.35-0.84** |
| **Parental understanding, (Ref: Low)** |  |  |  |  |
| High | 1.22 | 0.82-1.81 | 1.16 | 0.75-1.82 |
| **Parental respect, (Ref: Low)** |  |  |  |  |
| High | **0.64\*** | **0.43-0.97** | 0.67 | 0.36-1.24 |
| **Have close friends, (Ref: No)** |  |  |  |  |
| Yes | 1.20 | 0.68-2.12 | 1.08 | 0.48-2.40 |
| **Loneliness, (Ref: No)** |  |  |  |  |
| Yes | 1.08 | 0.70-1.68 | 0.93 | 0.48-1.83 |
| **Suicide attempt, (Ref: No)** |  |  |  |  |
| Yes | **1.75\*** | **1.08-2.83** | 1.49 | 0.83-2.68 |
| **Violence, (Ref: No)** |  |  |  |  |
| Yes | 1.20 | 0.55-2.59 | 1.06 | 0.48-2.34 |
| **Bullied, (Ref: No)** |  |  |  |  |
| Yes | **2.67\*\*** | **1.35-5.27** | **3.38\*\*** | **1.69-6.79** |
| **Sexual intercourse, (Ref: No)** |  |  |  |  |
| Yes | **3.05\*\*\*** | **1.71-5.44** | **2.97\*\*** | **1.46-6.03** |
| **Truancy, (Ref: No)** |  |  |  |  |
| Yes | **1.68\*** | **1.11-2.54** | 1.01 | 0.64-1.59 |
| **Drinking alcohol in the past 30 days, (Ref: No)** |  |  |  |  |
| Yes | **5.38\*\*\*** | **3.75-7.71** | **3.37\*\*\*** | **2.27-5.00** |
| **Sedentary behaviors, (Ref: No)** |  |  |  |  |
| Yes | **1.81\*\*** | **1.25-2.60** | **1.54\*** | **1.05-2.27** |
| **Low fruit/vegetables intake, (Ref: No)** |  |  |  |  |
| Yes | 0.66 | 0.30-1.47 | 1.01 | 0.33-3.08 |

\* $p < 0.05$ ; \*\* $p < 0.01$ ; \*\*\* $p < 0.001$

## Discussion

Tobacco smoking is one of the most dangerous risk behaviors. In this study, we used the data from the Vietnam GSHS 2013 and 2019 to describe the prevalence of tobacco smoking and associated factors among school students aged 13-17 years old. We found that the prevalence of traditional tobacco smoking decreased significantly from 2013 to 2019. The prevalence of students using e-cigarettes in the last 30 days was 2.6% in 2019. Factors associated with higher odds of traditional smoking included gender, mental health problems, sexual intercourse, truancy, and alcohol consumption. Risk factors of e-cigarettes smoking included gender, mental health problems, bullying, sexual intercourse, truancy, alcohol consumption, and sedentary behaviors. In contrast, the protective factor for traditional smoking was parental monitoring; for e-cigarettes smoking were parental monitoring and parental respect.

We found a significant decrease in the prevalence of traditional tobacco smoking among the students aged 13-17 years old in Vietnam from 2013 to 2019. This tendency can be explained by positive and effective activities of Vietnam steering committee on smoking and health and other partner organizations. The communicating and educating activities about tobacco harms were performed with many methods which was attracted attention and increasing knowledge of community. The Global Adult Tobacco Use Survey (GATS 2015) in Vietnam also indicated a decreasing trend in adult smoking rates, from 23.8% in 2010 to 22.5% in 2015 [19]. In the literature, the prevalence of tobacco smoking varies across countries and the time surveyed. According to a multi-country analysis of tobacco use among adolescents aged 13-15 years old (data of GSHS from 44 countries, 110 sites) [20], the overall prevalence of tobacco use ranged from 0.9% in Tajikistan (2006) to 32.8% in Chile (2005). In South-East Asia region, the prevalence of cigarette smoking ranged from 1.2% in India (2007) to 11.7% in Indonesia (2007). Within the Western Pacific region, the prevalence of tobacco use ranged from 3.5% in China (2003) to 12.3% in Philippines, Luzon (2003) [20].

Regarding the trend of tobacco use among the adolescents, several studies have also shown the declining trend. Study of Choi et al. using data from the Health and Nutrition Examination Survey from 1998 to 2013 and the Korea Youth Risk Behavior Web-based Survey from 2005 to 2013 indicated a clear decline trend among girls [21]. The global report on the trends of tobacco use from 2000 and projected to 2025 showed that the smoking prevalence among people aged 15 to 25 years will decrease from 22.6% in 2010 to 14.2% in 2025 [22].

In line with previous studies, our study confirmed that the risk factors of tobacco smoking included male gender [21, 23, 24], mental health problems [25, 26], having sexual intercourse [27], truancy [28, 29], and alcohol consumption [23, 24, 28].

The prevalence of e-cigarette use among youth widely varies across countries. For example, the prevalence of daily use of e-cigarette among New Zealander youth aged 14-15 in 2019 was 3.1% [30], 4.4% of adolescents aged 15-17 years in Taiwan reported ever having used e-cigarettes in 2015 [31], and very high in the US with 27.5% of high school students and 10.5% of middle school students reported current e-cigarette use (in the past 30 days) in 2019 [32]. In addition, the prevalence of e-cigarette use in youth increased rapidly over a short period of time in some countries [33]. Current e-cigarette use among US high school students, for instance, rose from 1.5% to 16% during the 2011-2015 period [34] before reaching the current level. The prevalence of e-cigarette uses among Vietnamese students in our study (2.6%) seemed to be lower than the above-mentioned countries. This can be explained that currently e-cigarettes are not allowed to be imported to Viet Nam, all the e-cigarette products in the markets are illegal/smuggled products.

Most e-cigarettes contain nicotine that can lead to respiratory ailments, negative impacts on attention, learning, and memory among adolescents and young adults, while long-term health effects of e-cigarette use remain unknown [35-37]. Moreover, some evidence indicates that e-cigarette use in adolescents can be a gateway to the initiation of drugs and other tobacco products use [37, 38], thereby causing more harmful effects. The design of e-cigarette devices and its variety of liquid solutions flavors (i.e., menthol, fruit, coffee, candy, etc.) that hide the harsh taste of nicotine can be an appeal to adolescents. Another concern is that many adolescents and young adults may perceive that vaping is cool, socially acceptable, or no harmful effects [39, 40], and adolescents are susceptible to peer influences, which can make e-cigarette use quickly become popular among adolescents. Therefore, despite the low prevalence of e-cigarette smoking in adolescents in Vietnam, it should be given adequate attention from experts working on tobacco control. Also, we found that e-cigarette use was associated with other unhealthy behaviors among Vietnamese students, such as alcohol drinking, sexual intercourse, truancy, and sedentary behaviors, suggesting that health education programs for adolescents should comprehensively target on this compound risk factors rather than individual factor. However, longitudinal studies are needed to determine causal factors associated with e-cigarette use among Vietnamese adolescents.

It is noticed that a high level of parental monitoring was associated with lower odds of smoking among the students. Vietnam is a country with a traditional family-oriented culture, parental factors always play a crucial role in children’s behaviors. Some studies have also shown the protective effects of parental factors on children’s behaviors, including smoking status.

Castrucci’s research about the association between parenting style and US adolescent smoking indicated that parenting style was associated with a 26% reduction in adolescent smoking [32]. A study in Cook Islands, Curacao, and East Timor also found that parental monitoring, which was measured in four aspects: understanding, knowing free time activities of children, (went over things without permission, and checking homework), related to the lower odds of smoking of adolescents aged 13 to 17 years old [41]. Together the results highlighted the need to incorporate parental involvement in programs that are designed to reduce tobacco smoking among adolescents.

## Limitations

There are several limitations of this study. First, despite a national representative school-based sampling frame, results from this study cannot be generalized to all Vietnamese adolescent as those who were not attending schools were not included in this study. In addition, due to the cross-sectional design, we were unable to determine causal factors of a decreased trend in cigarette smoking as well as the emerging of e-cigarette use among Vietnamese students. Third, data were self-reported and might be subject to recall and response bias.

## Data Availability

The datasets analyzed for the current study are not publicly available but are available upon reasonable request

---

**Supplemental Table 1:** Prevalence of traditional tobacco smoking by students’ characteristics.

| Factor | All students |  |  |  | Male students |  |  |  |
| --- | --- | --- | --- | --- | --- | --- | --- | --- |
|  | 2013 |  | 2019 |  | 2013 |  | 2019 |  |
|  | % | 95% CI | % | 95% CI | % | 95% CI | % | 95% CI |
| **Gender** |  |  |  |  |  |  |  |  |
| Male | 9.6 | 6.8–13.3 | 4.9 | 3.6–6.5 | -- | -- | -- | -- |
| Female | 1.7 | 0.9–2.9 | 1.0 | 0.7–1.5 | -- | -- | -- | -- |
| **Age** |  |  |  |  |  |  |  |  |
| 13 | 2.7 | 1.9–3.8 | 1.9 | 1.2–2.9 | 4.0 | 2.6-6.0 | 2.8 | 1.5-5.0 |
| 14 | 3.3 | 2.4–4.5 | 2.8 | 1.8–4.5 | 5.0 | 3.5-6.9 | 5.0 | 2.6-9.2 |
| 15 | 6.2 | 4.7–8.2 | 2.0 | 1.1–3.5 | 11.0 | 8.2-14.5 | 3.8 | 2.2-6.5 |
| 16 | 7.0 | 4.0–12.2 | 2.6 | 1.7–3.9 | 13.3 | 8.0-21.3 | 4.3 | 2.9-6.2 |
| 17 | 9.2 | 5.0–16.5 | 4.6 | 3.3–6.5 | 18.4 | 9.2-33.4 | 8.4 | 5.7-12.3 |
| **Parental monitoring** |  |  |  |  |  |  |  |  |
| Low | 7.2 | 4.9–10.4 | 3.3 | 2.4–4.5 | 13.1 | 8.3-20.0 | 6.0 | 4.1-8.8 |
| High | 3.3 | 2.3–4.9 | 2.1 | 1.4–3.2 | 6.1 | 4.1-9.1 | 3.6 | 2.5-5.3 |
| **Parental understanding** |  |  |  |  |  |  |  |  |
| Low | 5.9 | 4.1–8.3 | 3.1 | 2.3–4.3 | 10.6 | 7.1-15.5 | 5.1 | 3.3-7.8 |
| High | 4.1 | 2.9–5.9 | 2.5 | 1.7–3.7 | 7.4 | 5.1-10.7 | 4.7 | 3.2-7.0 |
| **Parental respect** |  |  |  |  |  |  |  |  |
| Low | 7.0 | 4.6–10.4 | 5.8 | 4.1-8.2 | 11.2 | 6.8-18.0 | 8.8 | 5.9-13.0 |
| High | 4.2 | 3.0–5.8 | 2.2 | 1.6-3.0 | 8.0 | 5.7-11.2 | 4.1 | 2.8-5.8 |
| **Have close friends** |  |  |  |  |  |  |  |  |
| No | 8.0 | 4.4–13.9 | 3.0 | 1.9–4.8 | 12.3 | 6.6-21.9 | 5.5 | 3.5-8.7 |
| Yes | 5.1 | 3.7–7.1 | 2.7 | 2.1–3.5 | 9.2 | 6.3-13.4 | 4.7 | 3.4-6.4 |
| **Loneliness** |  |  |  |  |  |  |  |  |
| No | 4.6 | 3.2–6.6 | 2.4 | 1.9–3.2 | 8.5 | 5.8-12.5 | 4.3 | 3.2-5.8 |
| Yes | 11.2 | 7.9–15.6 | 5.2 | 3.6–7.4 | 18.5 | 12.5-26.6 | 9.0 | 6.2-13.1 |
| **Suicide attempt** |  |  |  |  |  |  |  |  |
| No | 4.4 | 2.8–6.8 | 2.4 | 1.8–3.1 | 7.8 | 4.9-12.4 | 4.4 | 3.2-5.9 |
| Yes | 9.4 | 6.8–12.7 | 4.7 | 3.3–6.7 | 19.9 | 13.4-28.5 | 8.3 | 5.3-12.6 |
| **Violence** |  |  |  |  |  |  |  |  |
| No | 4.7 | 3.1–7.1 | 2.1 | 1.6–2.8 | 8.8 | 5.6-13.6 | 3.9 | 2.9-5.3 |
| Yes | 7.8 | 6.1–9.9 | 8.4 | 5.8–11.9 | 12.0 | 9.2-15.5 | 10.1 | 7.1-14.0 |
| **Bullied** |  |  |  |  |  |  |  |  |
| No | 5.0 | 3.3–7.7 | 2.5 | 1.9–3.3 | 8.6 | 5.3-13.7 | 4.5 | 3.3-6.1 |
| Yes | 6.0 | 4.5–8.0 | 7.7 | 4.7–12.4 | 12.0 | 8.9-16.0 | 10.3 | 6.2-16.8 |
| **Sexual intercourse** |  |  |  |  |  |  |  |  |
| No | 3.6 | 2.7–4.7 | 2.2 | 1.7–2.9 | 6.3 | 4.5-8.8 | 3.9 | 2.8-5.4 |
| Yes | 25.4 | 17.0–36.3 | 12.2 | 8.4–17.6 | 36.7 | 25.6-49.5 | 17.1 | 11.5-24.8 |
| **Truancy** |  |  |  |  |  |  |  |  |
| No | 3.4 | 2.5–4.6 | 2.0 | 1.4–2.7 | 5.9 | 4.2-8.4 | 3.7 | 2.5-5.5 |
| Yes | 13.4 | 9.3–18.7 | 6.9 | 5.5–8.8 | 21.0 | 14.2-29.8 | 9.5 | 7.0-12.9 |
| **Drinking alcohol in the past 30 days** |  |  |  |  |  |  |  |  |
| No | 2.0 | 1.3–3.0 | 1.2 | 0.8–1.7 | 3.8 | 2.3-6.3 | 2.2 | 1.5-3.3 |
| Yes | 15.5 | 11.8–20.1 | 8.3 | 6.4–10.9 | 21.3 | 15.9-27.9 | 13.0 | 9.3-17.7 |
| **Sedentary behaviors** |  |  |  |  |  |  |  |  |
| No | 4.3 | 3.1–6.1 | 2.5 | 1.7–3.7 | 7.9 | 5.3-11.6 | 4.6 | 3.1-6.7 |
| Yes | 6.4 | 4.7–8.8 | 3.2 | 2.4–4.3 | 11.2 | 8.3-15.0 | 5.4 | 3.7-7.9 |
| **Low fruit/vegetables intake** |  |  |  |  |  |  |  |  |
| No | 5.5 | 3.1–9.4 | 4.2 | 2.0–8.6 | 10.0 | 5.0-18.9 | 4.7 | 2.3-9.4 |
| Yes | 5.3 | 3.9–7.2 | 2.7 | 2.1–3.5 | 9.5 | 6.9-13.0 | 4.9 | 3.6-6.5 |

**Supplemental Table 2:** Prevalence of e-cigarettes smoking by students’ characteristics.

| Factor | All students |  | Male students |  |
| --- | --- | --- | --- | --- |
|  | % | 95% CI | % | 95% CI |
| **Gender** |  |  |  |  |
| Male | 3.6 | 2.7-4.8 | -- | -- |
| Female | 1.5 | 0.9-2.5 | -- | -- |
| **Age** |  |  |  |  |
| 13 | 3.1 | 1.5-6.2 | 3.8 | 1.5-9.1 |
| 14 | 1.8 | 1.0-3.0 | 2.1 | 1.2-3.5 |
| 15 | 1.9 | 1.3-3.0 | 3.0 | 1.8-4.9 |
| 16 | 2.5 | 1.6-3.8 | 3.6 | 2.3-5.5 |
| 17 | 3.9 | 2.4-6.2 | 6.7 | 4.1-10.8 |
| **Parental monitoring** |  |  |  |  |
| Low | 3.2 | 2.4-4.3 | 4.9 | 3.4-6.9 |
| High | 1.7 | 1.2-2.4 | 2.5 | 1.8-3.5 |
| **Parental understanding** |  |  |  |  |
| Low | 2.8 | 2.1-3.8 | 4.1 | 2.9-5.7 |
| High | 2.2 | 1.5-3.2 | 3.3 | 2.3-4.8 |
| **Parental respect** |  |  |  |  |
| Low | 5.1 | 3.4-7.7 | 6.5 | 3.9-10.6 |
| High | 2.1 | 1.5-2.8 | 3.1 | 2.2-4.3 |
| **Have close friends** |  |  |  |  |
| No | 3.4 | 2.1-5.5 | 4.8 | 2.6-8.7 |
| Yes | 2.4 | 1.8-3.3 | 3.5 | 2.6-4.7 |
| **Loneliness** |  |  |  |  |
| No | 2.1 | 1.5-3.0 | 3.2 | 2.3-4.5 |
| Yes | 5.0 | 3.5-7.1 | 6.7 | 4.4-10.2 |
| **Suicide attempt** |  |  |  |  |
| No | 2.0 | 1.4-2.7 | 3.1 | 2.2-4.2 |
| Yes | 5.3 | 3.6-7.7 | 7.4 | 4.7-11.5 |
| **Violence** |  |  |  |  |
| No | 2.1 | 1.4-3.0 | 3.2 | 2.3-4.3 |
| Yes | 6.2 | 4.5-8.5 | 6.2 | 4.2-9.0 |
| **Bullied** |  |  |  |  |
| No | 2.1 | 1.5-2.9 | 2.9 | 2.1-4.1 |
| Yes | 10.1 | 6.7-14.9 | 13.3 | 8.7-19.7 |
| **Sexual intercourse** |  |  |  |  |
| No | 2.1 | 1.5-2.8 | 2.9 | 2.2-4.0 |
| Yes | 10.6 | 6.6-16.7 | 13.2 | 7.6-21.9 |
| **Truancy** |  |  |  |  |
| No | 1.9 | 1.4-2.6 | 3.1 | 2.3-4.2 |
| Yes | 5.5 | 3.6-8.2 | 5.4 | 3.4-8.3 |
| **Drinking alcohol in the past 30 days** |  |  |  |  |
| No | 1.0 | 0.7-1.5 | 1.9 | 1.3-2.8 |
| Yes | 7.5 | 5.5-10.2 | 8.9 | 6.5-12.1 |
| **Sedentary behaviors** |  |  |  |  |
| No | 1.6 | 1.2-2.2 | 2.5 | 1.9-3.5 |
| Yes | 3.7 | 2.7-5.0 | 5.4 | 3.8-7.6 |
| **Low fruit/vegetables intake** |  |  |  |  |
| No | 3.7 | 1.8-7.1 | 3.1 | 1.1-8.4 |
| Yes | 2.4 | 1.8-3.3 | 3.7 | 2.7-4.9 |

